# A First Analysis of Excess Mortality in Switzerland in 2020

**DOI:** 10.1101/2021.03.14.21253551

**Authors:** Isabella Locatelli, Valentin Rousson

**Affiliations:** Center for Primary Care and Public Health (Unisanté), University of Lausanne, Switzerland, Route de Berne 113, CH-1010 Lausanne, Switzerland

**Keywords:** all-cause mortality, COVID-19, directly standardized mortality rate, life expectancy, Switzerland

## Abstract

**Objective:** To quantify the excess all-cause mortality in Switzerland in 2020, a key indicator for assessing direct and indirect consequences of the COVID-19 pandemic.

**Methods:** Using official data on deaths in Switzerland, all-cause mortality in 2020 was compared with that of previous years using directly standardized mortality rates, age- and sex-specific mortality rates, and life expectancy.

**Results:** The standardized mortality rate was 8.6% higher in 2020 than in 2019, returning to the level observed 5-6 years ago. This increase was greater for men (10.4%) than for women (7.0%), and was statistically significant only for men over 70 years of age, and for women over 75 years of age. The decrease in life expectancy in 2020 compared to 2019 was about 1%, with a loss of 9.4 months for men and 5.1 months for women.

**Conclusions:** There was an excess mortality in Switzerland in 2020, linked to the COVID-19 pandemic. However, as this excess only concerned the elderly, the resulting loss of life expectancy was restricted to a few months.

## Introduction

The year 2020 will be remembered as the year of the global spread of the COVID-19 pandemic caused by the acute respiratory syndrome coronavirus 2 (SARS-CoV-2). By early 2021, more than 100 million cases were confirmed worldwide and more than 2.5 million deaths were attributed to COVID-19. At a crossroads in the heart of Europe, after a first death recorded on March 5, 2020, Switzerland was particularly affected by the pandemic with more than 550’000 cases and 10’000 deaths attributed to COVID-19 (about 7400 in 2020) for a population of 8.7 million. In 2020, Switzerland had the 21st highest mortality rate due to COVID-19 among 156 countries with a population of at least one million (https://www.worldometers.info/coronavirus/).

Various restrictive and protective measures have been taken around the world, including in Switzerland, to fight the pandemic. The costs in terms of social burdens and economic consequences and benefits in terms of health of these measures are difficult to quantify and will be the subject of long discussions for many years. For such assessments, one key indicator is all-cause mortality. Unlike the specific mortality attributed to COVID-19, which may depend on partly unreliable diagnoses, all-cause mortality is a robust indicator that allows the direct and indirect effects of a pandemic to be taken into account.

In this paper, we propose an analysis of all-cause mortality in Switzerland for the year 2020, with the aim of quantifying the excess mortality observed in 2020 compared to previous years in one of the countries with the highest life expectancy in the world (second behind Japan according to the World Health Organization, 2016). In Switzerland, the Federal Statistical Office (FSO) has been updating weekly the number of deaths by sex and age group for each of the 53 weeks of the year 2020, making these figures available to the public. Since it usually takes a few years to obtain complete and consolidated statistics on deaths in a country, our analysis is referred to as a “first analysis”. This is also the reason why few such analyses have been conducted and published so far worldwide for the year 2020. For example, Aburto et al (2021) studied all-cause mortality in England and Wales, while Andrasafay and Goldman (2021) did so in the United States, but both studies were not based on the entire year 2020. Given the potential importance of such analyses in the current (urgent) scientific and political debates on the COVID-19 pandemic, we believe that even results that are not entirely definitive deserve to be presented and published.

When analyzing and comparing mortality over the years, a simple look at the total number of deaths is usually misleading because of the tendency of the population to increase, which naturally implies an increasing number of deaths. Therefore, the number of deaths must be divided by the population size, resulting in a so-called crude mortality rate. However, the latter still does not account for changes in the age (and possibly sex) distribution of the population over time (Curtin and Kline, 1995). Indeed, the age structure of the European and Swiss population changes each year, moving towards an increasingly older population (FSO, Age Structure of Switzerland, 1860-2050). As the population ages, crude mortality rates will stay constant even as age-specific mortality decreases.

Standardized (or adjusted) mortality rates were developed in the first half of the 19th century (Neison, 1844) as a summary measure of mortality that controls for the changes in the age and sex distribution of a population. With direct standardization, which is perhaps the simplest method of adjustment, the age- and sex-specific mortality rates of a study population are applied to the age and sex distribution of a standard population, resulting in the all-cause mortality that would be observed if the age- and sex-specific mortality rates of the study population were applied to the age and sex structure of the standard (Fleiss, 2003). One can then compare the mortality observed in two different years (such as 2020 and 2019) via a ratio of two standardized mortality rates, as suggested for example by Miettinen (1972).

To quantify the excess mortality observed in Switzerland in the year 2020, we will therefore calculate and compare the standardized mortality rates over the years using the Swiss population at the beginning of the year 2020 as the standard. We will then make similar comparisons in selected age groups, separately for men and women, in order to identify the specific ages most affected by excess mortality. Finally, we will also calculate and compare the life expectancies that would result from the age- and sex-specific mortality rates observed over the years, placing the life expectancy of 2020 in a secular perspective.

## Data

We used official data on deaths in Switzerland by sex and age group published for 2020 and previous years (going back to 1970) by the Swiss Federal Statistical Office, FSO (https://www.bfs.admin.ch/bfs/fr/home/statistiques/population/naissances-deces/deces.html).

Our last access to this site was on March 9, 2021. The number of deaths, separately for men and women, was available by 1-year age groups (with a last open class of 110+) until 2019, and by 5-year age groups (with a last open class of 90+) for the year 2020, for which the number of deaths is provided for each week. By convention, the year 2020 is divided into 53 weeks, the first of which begins on December 30, 2019 and the last of which ends on January 3, 2021. We therefore corrected the number of deaths in these two weeks that spill over into two other years by excluding 2/7 of the deaths in the first and 3/7 in the last. For leap years (including 2020), we further excluded 1/366 of the deaths.

The size of the Swiss population as of January 1, 2011-2020, stratified by sex and by one-year age groups (with a last open class of 100+), can be found in the FSO database (https://www.bfs.admin.ch/bfs/fr/home/statistiques/population/effectif-evolution.html) and for January 1, 1876-2019, in the Human Mortality Database, HMD (https://www.mortality.org/). Since the differences for the overlapping years are negligible, we considered the HMD source for the years 1970-2010 and the FSO source for the years 2011-2020. Life expectancy for both sexes (and pooled) is also available in the HMD database for the period 1876-2018.

## Methods

Directly standardized mortality rates (dSMR) were used to compare mortality in 2020 with mortality in previous years, always considering 2020 as the standard. Algebraically, a dSMR is an average of the age- and sex-specific mortality rates observed in a given year, weighted by the age and sex distribution of the standard population. The dSMR for year *y* (*y* = 1970, …. 2019) is therefore defined as follows:

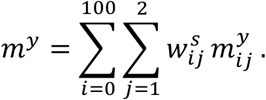

In this formula, 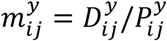represents the age- and sex-specific mortality rate for age *i* (*i* = 0,1, …, 99,100 +) and sex *j* (*j* = 1 for men and *j* = 2 for women) in year *y* (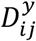 being the number of deaths and 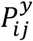 the size of the population for age *i* and sex *j* in year *y*), while 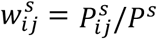 is the proportion of people with age *i* and sex *j* in the reference year *s, P*^s^ being the total size of the population in that year. The dSMR for the reference year *y* = *s* = 2020 corresponds to the crude mortality rate in that year, so that *m*^2020^ = *D*^2020^/*P*^2020^, *D*^2020^ being the total number of deaths observed in year 2020.

In order to evaluate an excess mortality in 2020, we compared *m*^2020^ with *m*^y^ for various years *y* via a relative change in dSMR (expressed in %) calculated as follows:

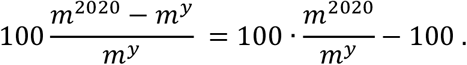

Although these quantities were obtained on the whole Swiss population, it is still important to calculate confidence intervals around them since “the number of events that actually occurred may be considered as one of a large series of possible results that could have arisen under the same circumstances’’ (Curtin and Klein 1995). A 95% confidence interval for a relative change can be obtained as follows (Breslow and Day 1987, Fay 1999):

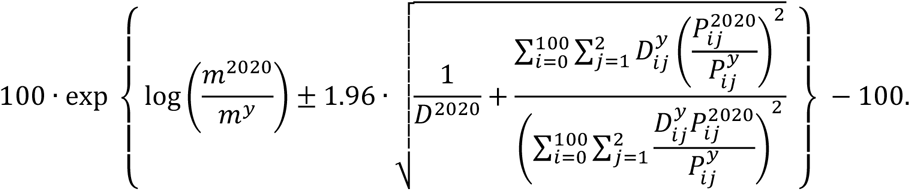

A relative change was considered statistically significant when the corresponding 95% confidence interval did not contain the value 0. Similar calculations were made for different sub-populations, such as men and women, or for selected age groups, although we could not consider finer groups than those available for the number of deaths in 2020 (5-year age groups with a last open class of 90+). In order to get smoother results that did not depend too much on what might have happened in a given year, we also carried out comparisons of the dMSR observed in 2020 with the dMSR obtained by pooling data from several previous years.

Another way to summarize mortality in a given year *y* is to calculate the life expectancy at birth *LE*^y^ resulting from the age- and sex-specific mortality rates observed in that year. To calculate *LE*^y^ for a given year *y* = 1970, …. 2020, we applied a piecewise exponential model (Friedman, 1982) to the age and sex-specific mortality rates observed in that year, using the age groups available in 2020. We validated our method by comparing the resulting life expectancies for the years 1970-2018 with those available in the HMD database, finding an average absolute difference of 0.07%, with no difference exceeding 0.3% (or even 0.08% since 2010). Life expectancies for the years 1876-1969 were taken directly from the HMD database. We were then able to compare *LE*^2020^ with *LE*^y^ for various years *y*, either via an absolute change in life expectancy (expressed in months), 12(*LE*^2020^ − *LE*^y^), or via a relative change in life expectancy (expressed in %), 100(*LE*^2020^ − *LE*^y^)/*LE*^y^.

## Results

Figure 1 shows the crude and standardized mortality rates in Switzerland calculated for the years 1970-2020. In contrast to the crude mortality rates, which have remained roughly constant, the standardized mortality rates have decreased by a factor of about 2.5 over the last 50 years, reflecting the continuous and impressive reduction in mortality. In 2020, however, the standardized mortality rate was higher than in previous years, although it is still lower than in 2015 and in all years before (including) 2013. Similar conclusions were drawn when calculations were made separately for men and women (Supplementary Figure 1).

**Figure 1.**
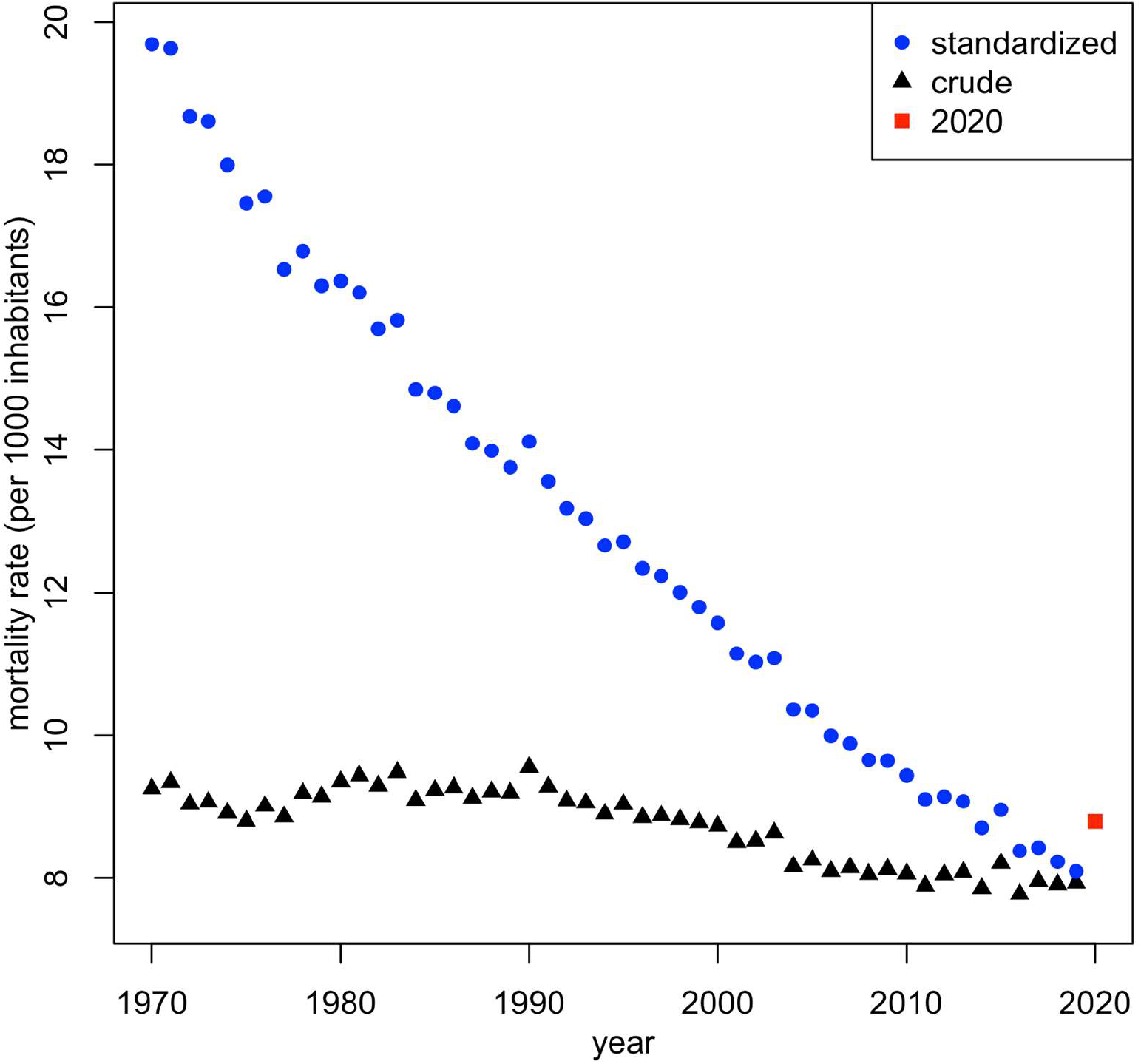
Crude and directly standardized mortality rates in Switzerland for the period 1970-2020 (data from the Swiss Federal Statistical Office and the Human Mortality Database).

Table 1 presents the relative changes in dSMR when comparing the year 2020 with the years 2019, 2015 and 2010, and with pooled years 2015-2019 and 2010-2019. One can see that the observed mortality in 2020 has increased by 8.6% compared to 2019, this relative difference being greater for men (10.4%) than for women (7.0%). In absolute numbers, this represents an increase of 7877 deaths (6013 after standardization) compared to 2019. Mortality has increased by 4.6% compared to years 2015-2019, and only by 0.8% compared to years 2010-2019, while it has decreased by 1.9% compared to 2015, and by 6.8% compared to 2010.

**Table 1:** Summary of changes in directly standardized mortality rates (dSMR) and in life expectancy comparing 2020 to previous years.

|  | dSMR |  |  | life expectancy |  |  |  |  |  |
| --- | --- | --- | --- | --- | --- | --- | --- | --- | --- |
| *change in*<br>*2020 vs:* | total | men | women | total |  | men |  | women |  |
|  | % | % | % | % | months | % | months | % | months |
| **2019** | **+8.6** | +10.4 | +7.0 | **-0.7** | **-7.3** | -1.0 | -9.4 | -0.5 | -5.1 |
| **2015-2019** | **+4.6** | +5.6 | +3.7 | **-0.3** | **-2.8** | -0.4 | -3.9 | -0.2 | -1.8 |
| **2010-2019** | **+0.8** | +0.9 | +0.7 | **+0.2** | **+2.0** | +0.2 | +2.1 | +0.2 | +1.8 |
| **2015** | **-1.9** | -1.7 | -2.0 | **+0.4** | **+3.9** | +0.5 | +4.8 | +0.3 | +3.0 |
| **2010** | **-6.8** | -8.4 | -5.3 | **+1.2** | **+11.6** | +1.4 | +13.2 | +1.0 | +10.1 |

Figure 2 shows the relative change in dSMR in 2020 compared to years 2019, 2015 and 2010, separately for specific age groups, and for men and women, together with 95% confidence intervals. One can see that the dSMR increased significantly in 2020 compared to 2019 for all age groups over 70 years for men, respectively for all age groups over 75 years for women. For younger age groups, we found no significant change in dSMR. Compared to 2015, the dSMR increased significantly in only one age group for men (85-90), and in no age group for women, while it decreased significantly in 5 age groups for men (between 40 and 75), and in 2 age groups for women (50-60 and 65-70). Compared to 2010, the dSMR decreased significantly in most age groups for both men and women, showing that the mortality in Switzerland was lower in 2020 than in 2010 at all ages, including in the oldest age groups.

**Figure 2.**
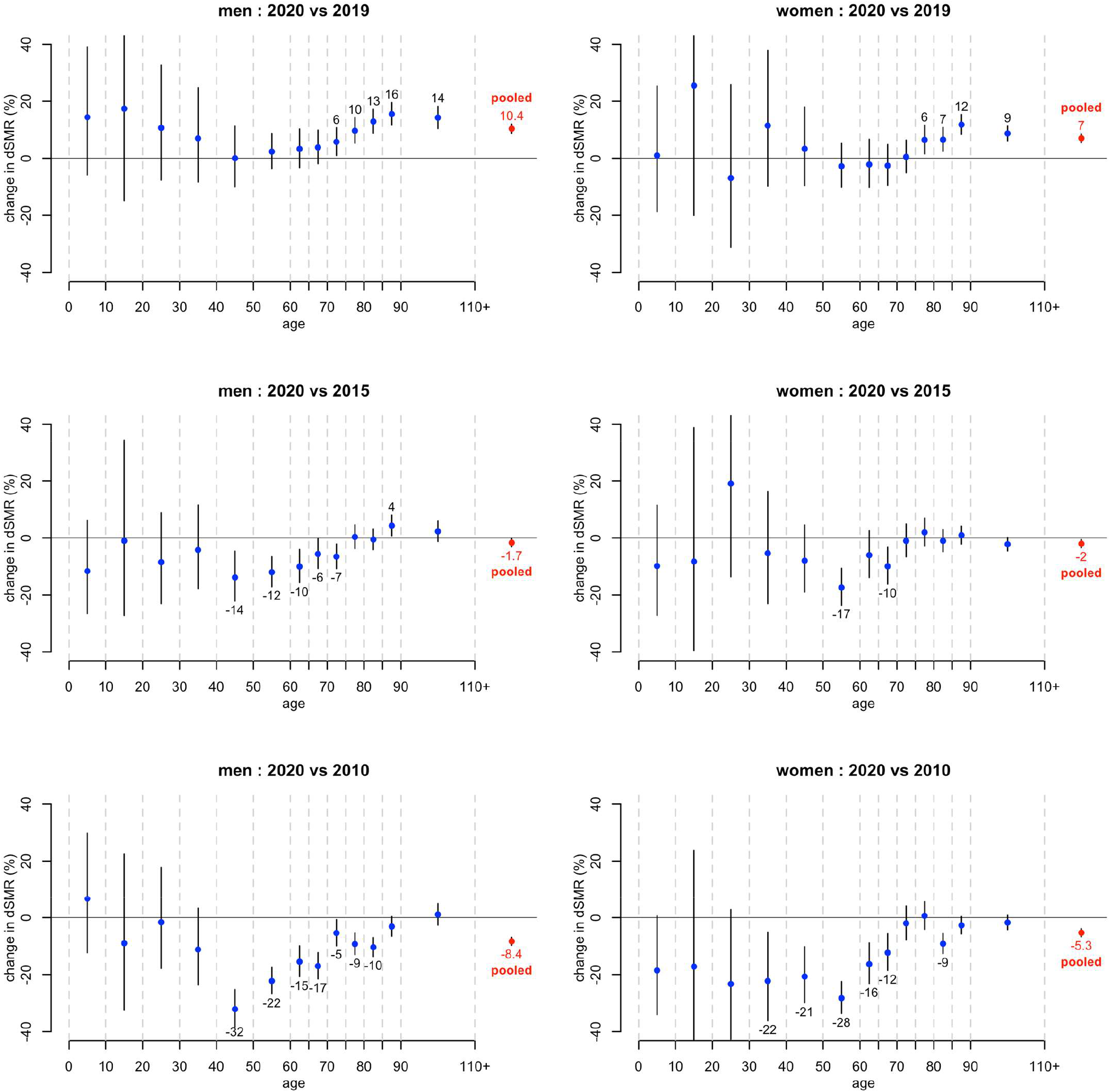
Relative change in directly standardized mortality rates (dSMR) when comparing 2020 with 2019, 2015 and 2010 in selected age groups, separately for men and women, as well as for pooled age classes, together with 95% confidence intervals. Estimated changes (expressed in %) are printed when statistically significant.

Figure 3 shows life expectancy at birth in Switzerland for the years 1876-2020, separately for men and for women. The secular increase of life expectancy is spectacular, rising from about 40 years in 1876, to 81.9 years for men and 85.6 years for women in 2019. Also spectacular is the sudden fall in life expectancy in 1918 following the Spanish Flu, where men lost 10.4 years and women 8.5 years compared to 1917. In 2020, life expectancy has also decreased compared to 2019, although the decrease was much lower, men reaching 81.1 years and women 85.2 years in 2020. Thus, the decrease was 9.4 months (or 1.0%) for men and 5.1 months (or 0.5%) for women. Averaged over both sexes, the decrease was 7.3 months (or 0.7%). See Table 1 for a comparison of life expectancy in 2020 with other years.

**Figure 3.**
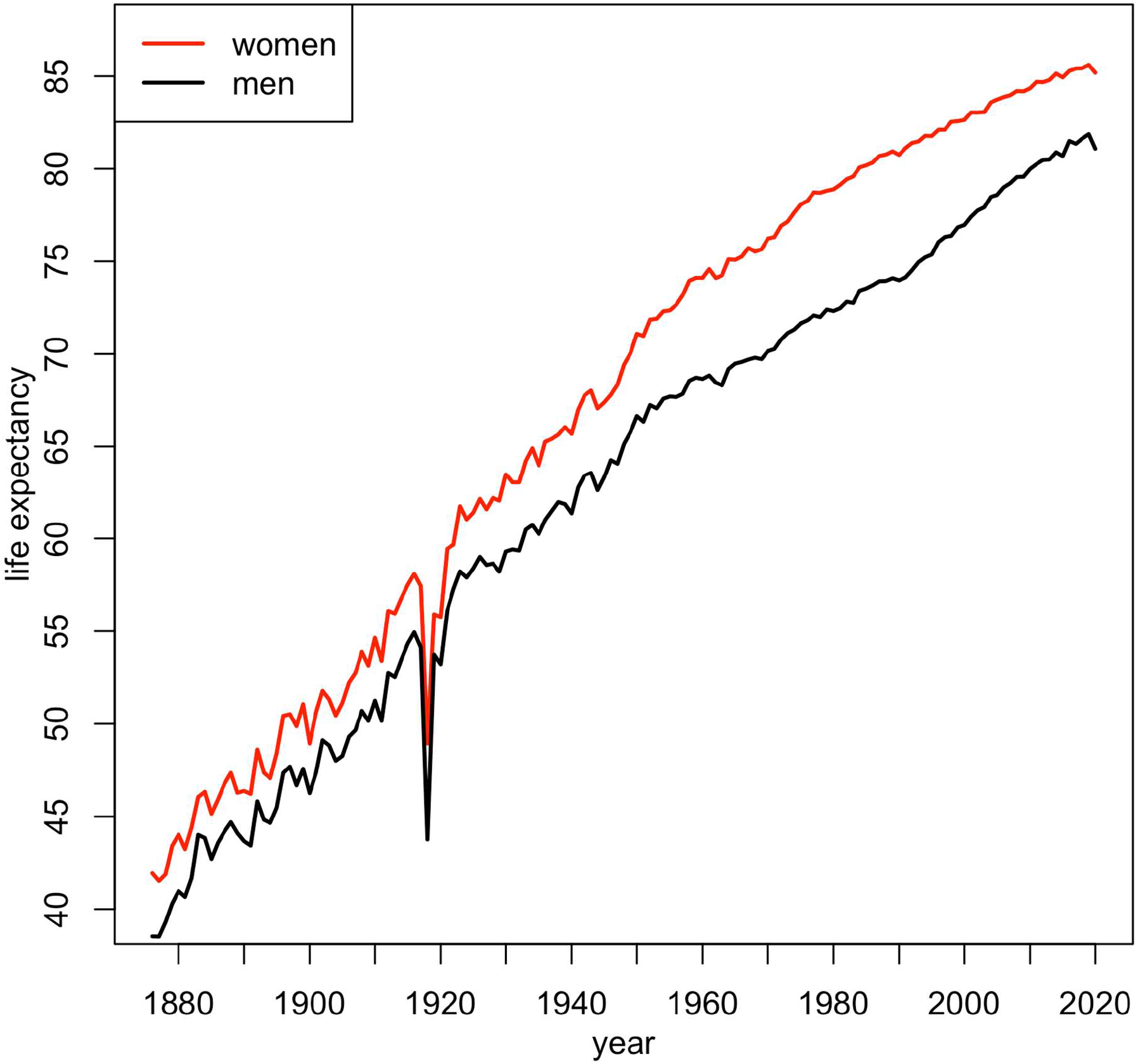
Life expectancy in Switzerland for the period 1876-2020 for men and women (data from the Swiss Federal Statistical Office and the Human Mortality Database).

## Discussion

In this paper, we have compared mortality in Switzerland in 2020 with that of previous years. We found that the standardized mortality rate is 8.6% higher in 2020 than in 2019, returning to the level observed 5-6 years ago. This increase is greater for men than for women, and is statistically significant only for the older age groups (over 70 years for men and over 75 years for women). The fact that the older age groups are the most affected explains why the decrease in life expectancy in 2020 compared to 2019 is much more modest (about 1%).

Standardization is a well-recognized technique in the field of mortality, making it possible to compare different populations or analyze the evolution of mortality over time, adjusting for differences or changes in the age structure of the population. Different standardization techniques have been developed and used, including direct and indirect standardization (see Fleiss 2003 for a discussion on the advantages and drawbacks of both methods). We have chosen to refer to direct standardization, primarily due to its intuitive nature. In addition, we particularly appreciate the interpretation of the ratio of directly standardized rates as the effect of exposure (in our case, year 2020) on mortality, net of the effect of changes in the age structure of the population (Miettinen, 1972). Finally, we note that comparisons made by direct standardization are relatively independent of the choice of the reference population, as shown by Spiegelman and Marks (1966).

Of course, as Woolsey (1959) and Evelback (1966) pointed out, the analysis of changes in standardized mortality rates does not entirely substitute the detailed analysis of changes in mortality rates by age. For example, when age-specific rates vary in opposite directions in different age groups, any summary measure, such as the standardized rate ratio, will mask these differences by providing a summary in which positive and negative variations in age-specific rates offset each other. This is why we have also carried out comparisons in selected age groups in order to identify the ages primary affected by changes in mortality.

In contrast to a standardized mortality rate, which is affected by mortality at all ages more or less equally, life expectancy at birth is a summary indicator of mortality that gives much more weight to mortality at a young age than at an advanced age. It is a popular indicator which is usually used to compare mortality across countries or periods. The loss of life expectancy between 2020 and 2019 (9.4 months for men and 5.1 months for women) is the greatest since 1944 for men (where the loss was 11.5 months), and since 1962 for women (where the loss was 5.8 months). In particular, it is greater than the loss observed in 2015, a year with a severe flu in winter and a heat wave in summer, responsible for a loss of 2.5 months for each sex.

On the other hand, the loss of life expectancy in 2020 in Switzerland is smaller than the loss of 1.1 years (13.6 months) in the United States, estimated by Andrasafay and Goldman (2021) based on the first 40 weeks of 2020, and of 1.2 years (14.4 months) for men and 0.9 years (10.8 months) for women, estimated by Aburto et al (2021) in England and Wales based on the first 47 weeks of 2020. This is consistent with the fact that the United States and the United Kingdom are among the countries that suffered the most from COVID-19 in 2020, reaching respectively the 8th and 11th highest mortality rate, with Switzerland ranked 21st. All these losses, however, are still much lower than those observed in 1918, the year of the Spanish Flu, when life expectancy decreased by more than 10 years for men and more than 8 years for women in Switzerland. Unlike COVID-19, the Spanish Flu mainly killed young adults between 20 and 40 years of age much more than the elderly.

Just as the massive increase in mortality observed in 1918 was mainly attributed to Spanish Flu, any excess mortality observed in 2020 will inevitably be interpreted as consequences (direct and indirect) of the COVID-19 pandemic, unquestionably the major event in 2020. In fact, the number of deaths attributed to COVID-19 in 2020 roughly corresponds to the number of additional all-cause deaths observed in 2020, involving both a majority of men and mainly people over 80 years of age.

As mentioned in the introduction, our analysis is not definitive because it takes some time before complete mortality data are available. This analysis is therefore intended to be repeated in a few months or years. In particular, it is possible than the mortality reported in 2020 will increase slightly in the coming months, as a couple of deaths might be reported with delay. We recognize that this is a limitation of our study. Nevertheless, having followed the weekly updates of the FSO figures since the very beginning of 2021, we expect that our results are not far from having converged.

Although it is somewhat too early to draw definitive conclusions, there is no doubt that there has been an excess mortality in Switzerland in 2020, linked to the COVID-19 pandemic and due to an increased mortality among the elderly. It is also clear that the decrease in life expectancy of about 1% is much less impressive than the increase in mortality of 8.6%. Of course, these results were achieved under important protection measures taken in 2020 by the government to fight the COVID-19 pandemic and it is very difficult to guess what would have happened without them. These measures, by reducing the number of contacts among people, have certainly limited the spread of the virus, and thus hospital overcrowding and mortality. On the other hand, the social and economic costs of these measures are also undeniable and there will be a lively debate to take stock of the actions undertaken. We believe that the loss of life expectancy observed in the pandemic year 2020, which we estimated at a few months, will be an important element in this debate.

## Data Availability

All data are publicy available

---

**Supplementary Figure 1.**
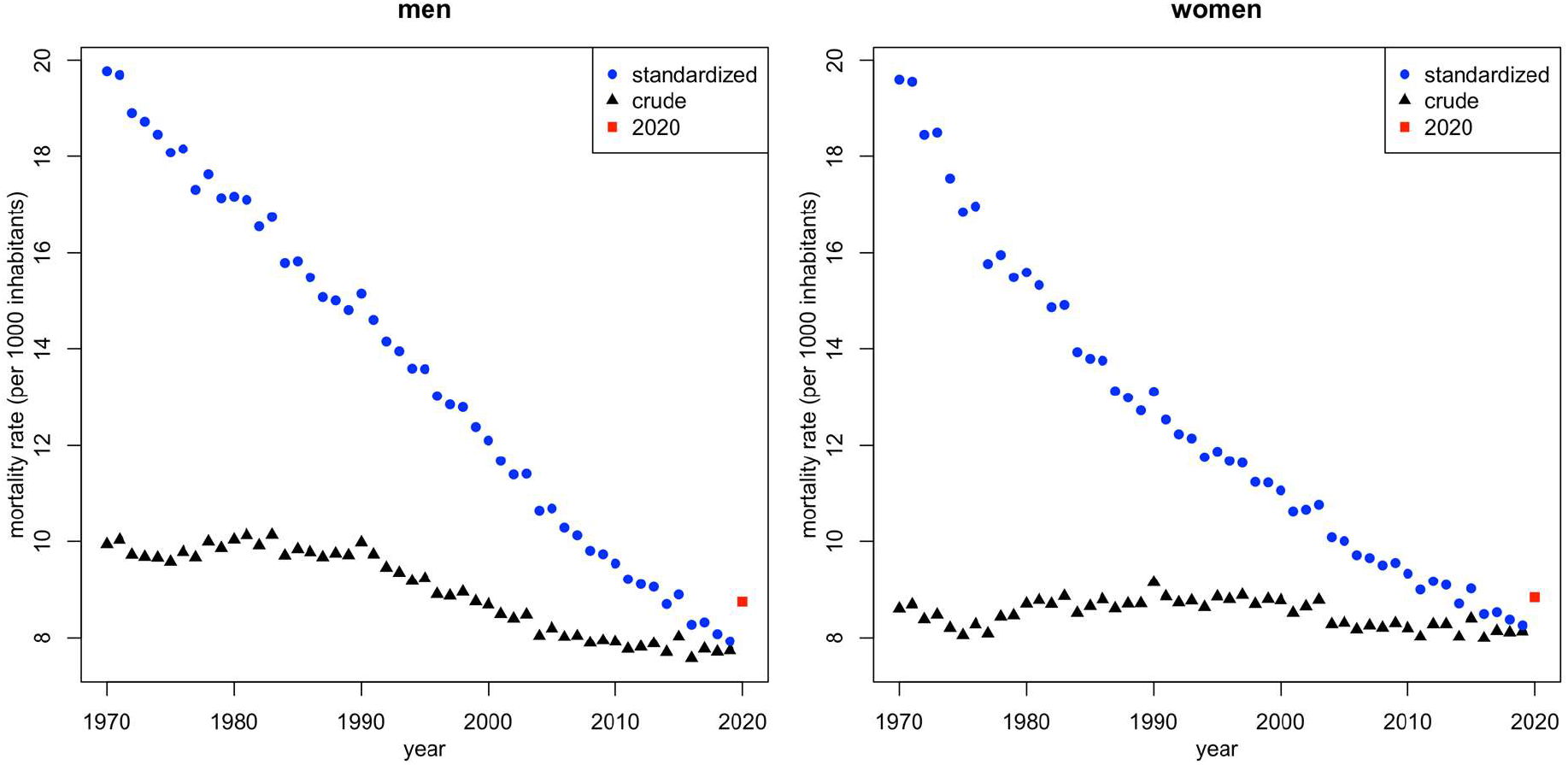
Crude and directly standardized mortality rates for men and women in Switzerland for the period 1970-2020 (data from the Swiss Federal Statistical Office and the Human Mortality Database).

## References

1. Aburto JM, Kashyap R, Schöley J, Angus C, Ermisch J, Mills MC, Beam Dowd J (2021). Estimating the burden of the COVID-19 pandemic on mortality, life expectancy and lifespan inequalities in England and Wales: A population-level analysis. Journal of Epidemiology and Community Health. Epub ahead of print.

2. Andrasafay T, Goldman N (2021). Reduction in 2020 US life expectancy due to COVID-19 and the disproportionate impact on the Black and Latino populations. PNAS, 118, e2014746118.

3. Breslow NE and Day NE (1987). Statistical methods in cancer research, Volume II – The design and analysis of cohort studies. London: Oxford University Press.

4. Curtin LR, Klein RJ (1995). Direct standardization (age-adjusted death rates). Healthy People 2000 Statistical Notes, 6, 1–10.

5. Elveback LR (1966). Discussion of “Indices mortality and tests of their statistical significance.” Human Biology, 38, 322–324.

6. Fay MP (1999). Approximate confidence intervals for rate ratios from directly standardized rates with sparse data. Comm. Statist. 28, 2141–2160

7. Fleiss JL, Levin B, Cho Paik M (2003). Chapter 19 in Wiley Series in Probability and Statistics Eds. Statistical methods for rates and proportions.

8. Friedman M (1982). Piecewise exponential models for survival data with covariates. Annals of Statistics, 10, 101–113.

9. Miettienen OS (1972) Components of the crude risk ratio. American Journal of Epidemiology, 96, 168–172.

10. Neison FGP (1844). On a method recently proposed for conducting inquiries into the comparative sanatory condition of various districts. Journal of the Royal Statistical Society of London (now the Royal Statistical Society), 7, 40–68

11. Spiegelman M and Marks HH (1966). Empirical testing of standards for the age adjustment of death rates by the direct method. Human Biology, 38, 280–292

12. Woolsey TD (1959). Adjusted death rates and other indices of mortality. Chapter 4 in F.E. Linder and R.D. Grove (Eds.), Vital statistics rates in the United States, 1900-1940. Washington, D.C.: U.S. Government Printing Office.

13. World Health Organization (2016). World Health Statistics 2016: Monitoring health for the SDGs Annex B: tables of health statistics by country, WHO region and globally. https://www.who.int/gho/publications/world_health_statistics/2016/Annex_B/en/.

